# A community-based mentoring scheme for pregnant and parenting adolescents in Sierra Leone: protocol for a hybrid pilot cluster randomised controlled trial

**DOI:** 10.1101/2023.11.05.23298118

**Authors:** C Fernandez Turienzo, M Kamara, L November, P Kamara, AM Kingsford, A Ridout, S Thomas, PT Seed, AH Shennan, J Sandall, PT Williams

**Author notes:** Corresponding author: Dr Cristina Fernandez Turienzo, Department of Women & Children’s Health, School of Life Course and Population Sciences, Faculty of Life Sciences & Medicine, King’s College London, Westminster Bridge Road, London, SE1 7EH, United Kingdom. Joint first authors (C Fernandez Turienzo, M Kamara, L November). Joint last authors (J Sandall, PT Williams).

## Abstract

**Background:** Sierra Leone has a very high maternal mortality rate, and this burden falls heavily on adolescents, a particularly vulnerable group; this is usually driven by poverty, lack of education and employment opportunities. In 2017, a local grassroots organisation, Lifeline Nehemiah Projects, developed a community-based mentoring intervention ‘2YoungLives’ (2YLs) for adolescent girls in Eastern Freetown. We aim to formally assess the feasibility and implementation of the 2YL mentorship scheme in new communities in Sierra Leone.

**Methods:** A hybrid type 2 pilot cluster randomised controlled trial of the 2YL mentoring scheme in urban and rural communities living around twelve peripheral health units (PHU) across five districts in Sierra Leone. Clusters will be matched into pairs and randomisation will be determined by computer-generated random numbers via a secure web-based system hosted by MedSciNet. All under-eighteen adolescents identified as pregnant in the community and/or the PHU are included. Feasibility (recruitment, retention, and attrition rates; data collection and completeness; sample calculation) and primary clinical outcome data (composite of maternal deaths, stillbirths, neonatal deaths) will be collected. A mixed-methods process evaluation will explore implementation outcomes, mechanisms of change, contextual factors, experiences of care, and health and wellbeing. A concurrent cost-consequence analysis will be undertaken. Main trial analysis will be pragmatic, by intention to treat, and a complementary per protocol analysis will also be included.

**Discussion:** Improving health and wellbeing for adolescent girls (including sexual and reproductive health) remains a top priority in Sierra Leone indicated by several government policies targeted to this group, in which maternal and infant mortality are still persistently high. Supporting these girls and facilitating their wellbeing is imperative, along with sensitisation of communities, strengthening of youth friendly services and collaboration with stakeholders at all levels (government, regional, community, family). We believe 2YL supports the global holistic agenda to integrate and implement interventions across health, education, and social systems in order to protect, nurture, and support the health and development potential of every adolescent girl, and thus become a model of good practice for adolescent pregnancy, to be adopted more widely in Sierra Leone and elsewhere.

**Trial registration:** ISRCTN registry ISRCTN32414369. Prospectively registered on 14/03/2022.

## Introduction

Maternal mortality rate in Sierra Leone remains very high despite recent improvements (717 deaths per 100 000 livebirths in 2019) [1] with adolescent pregnancy a leading cause of death for mothers; adolescent mothers are 40-60% more likely to die during childbirth [2,3]. A household survey conducted post-Ebola in 2015 indicated a maternal death rate of 1 in 10 for under-18-year-olds in a poor suburb of Eastern Freetown. A subsequent study examined the causes of this high incidence of maternal death in adolescents and highlighted intersecting health and socio-economic vulnerabilities [4]. Among key findings were that pregnant adolescents were often neglected by their families, particularly when being cared for by a non-parental adult, often sleeping on bare ground without a mosquito net, and fed once a day in exchange for heavy domestic duties such as water collection and laundering, in many cases with little protection from ongoing gender-based violence. This lack of adult care or support often leads to delays in care-seeking or complete lack of antenatal and delivery care, putting girls at high risk of death from undetected pre-eclampsia, untreated infections, anaemia, lack of birth preparation, and other common obstetric risks [4]. To achieve the Sustainable Development Goals target of reducing the maternal mortality rate to less than 70 per 100 000 live births by 2030, adolescent girls are a priority group [5–7].

These findings led a grassroots organisation, Lifeline Nehemiah Projects (LNP), to develop a holistic and locally designed community-based mentoring intervention, 2YoungLives (2YL), from pregnancy through to one-year post-birth for adolescent girls [8]. Mentors support mentees to take up antenatal care and hospital birth, re-establish family connections where this is safe and appropriate, run a small business, and return to education or start vocational training. They promote and model early health-seeking behaviour, and provide practical advice about childbirth, parenting and contraception. The 2YL scheme was developed in 2017 in a suburb of the capital, Freetown, and expanded to five new sites in 2018, 2019 and 2021. A formative evaluation compared outcomes of young girls pre-intervention with those of other women post 2YL intervention [9]. In the 2YL cohort of young women who had mentors there were no maternal deaths (0% vs 10%) and lower levels of perinatal mortality (6% vs 16%), and infant mortality (11% vs 26%). Nearly all women receiving the 2YL intervention gave birth with a skilled birth attendant and breastfed for longer than 6 months; and more than two thirds were using contraception by the baby’s first birthday, with no second pregnancies. All of the 2YL young women successfully ran a small business with support from their mentor, allowing them to learn business skills, eat well throughout pregnancy and provide for their babies. They reported increased self-confidence, supportive peer relationships, and a high level of satisfaction with the mentoring scheme [9]. Subsequent anecdotal evidence is also showing that the community engagement strategy pursued as an integral part of the early implementation is influencing widely-held ingrained attitudes towards practices which perpetuate gender inequality such as school non-attendance in pregnancy and child marriage.

Based on this preliminary data, the 2YL mentoring scheme is promising [10]. A theory of change was co-developed to understand how and why the 2YL intervention may work in adolescent girls in Sierra Leone. 2YL has potential to improve the health and wellbeing of girls and their babies and sustainably improve livelihoods. Relationship building, engagement and advocacy, educational, social, and economic empowerment, and respectful community engagement and involvement are important mechanisms of action to consider. However, more robust evidence is needed to understand the impact and mechanisms of how 2YL can address determinants of adolescent maternal mortality, and general health and wellbeing [10]. The overall aim of this study is therefore to assess the feasibility and implementation of the 2YL mentorship scheme in new communities in Sierra Leone to inform procedures for a subsequent fully powered cluster trial evaluating the effectiveness and social and economic impact of 2YL. Specifically, the study objectives include:

- To assess the integrity of study protocol including: recruitment, randomization procedure, data collection, retention procedures, acceptability, primary outcome measure to inform the sample size calculation, refinement of the intervention and training and supervision procedures.
- To assess pregnancy outcomes of adolescents receiving the 2YL intervention compared to those receiving usual care.
- To compare experiences of care, mentoring, health and wellbeing, and thriving among young women receiving the 2YL intervention compared to those receiving usual care.
- To evaluate the implementation, contextual factors and mechanisms of change of the 2YL intervention in order to understand the results and its potential impact.

## Methods

### Setting and study design

Sierra Leone, a low-income West African country with a population of 8.8 million, has a very high maternal mortality rate and a high burden of communicable and non-communicable diseases. It struggles to achieve basic universal health coverage and access to potable water, sanitation and hygiene [11]. Sexual violence and rape are common, and capacity to provide treatment to affected women and girls is extremely limited (State of Emergency over rape and sexual violence was declared in 2019) [12]. Sierra Leone also suffers an inadequacy of human resources for health; it has 2 skilled providers (doctors, nurses and midwives) per 10,000 population which is well below WHO’s health workforce targets for UHC and Sustainable Development Goal of 23 [13]. Suboptimal maternal and child health care contributes to premature deaths, disability, and devastating spending in a country with an incipient financial crisis exacerbated by the 2014 Ebola epidemic (and further weakened during the ongoing COVID-19 epidemic and the global fuel and food instability in the wake of the war in Ukraine). Despite recent improvements, adolescents account for nearly half of all maternal deaths, and neonatal, infant and child mortality are also high; infants of mothers who die are up to 10 times more likely to die within their first two years, while infants born to adolescent mothers are also at increased risk of sickness and death [1,2]. Although healthcare services are free for pregnant women, lactating mothers, and children below the age of five, antenatal attendance, access to skilled birth attendants and facility deliveries are low, and quality service delivery is often poor with disparities in access and availability to appropriate assessment and intervention, with delays in delivery, escalation of care and emergency maternity care [14].

The 2YL trial will run in the communities served by twelve peripheral healthcare units (PHUs); the clusters, representing a range of urban and rural settings in five of the administrative regions or districts of Sierra Leone. The design is a pragmatic hybrid pilot cluster-randomised controlled trial (cRCT) of the introduction of the 2YL mentoring adjunct to maternity care in Sierra Leone. The trial structure is based on the Medical Research Council’s (MRC’s) guideline for developing and evaluating complex interventions [15], and Curran and colleagues’ hybrid type 2 effectiveness-implementation research design that place similar focus on assessing effectiveness of an intervention as well as how best to implement it [16]. The trial will be reported according to the Consolidated Standards of Reporting Trials (CONSORT) statement for randomised pilot and feasibility trials [17], the Standard Protocol Items: Recommendations for Interventional Trials (SPIRIT) statement [18] (S1 SPIRIT Checklist) as well as the Template for Intervention Description and Replication (TIDieR) checklist for intervention description [19]. The trial has been prospectively registered in the International Standard Randomised Controlled Trial Number Registry (ISRCTN32414369) and approved by ethics committee at King’s College London UK (HR/DP-21/22-26320) and the Office of the Sierra Leone Ethics and Scientific Review Committee. The (SPIRIT) Figure adapted for the 2YL trial is shown below in Figure 1. Any protocol amendments will be notified to ethics committees and all collaborators informed.

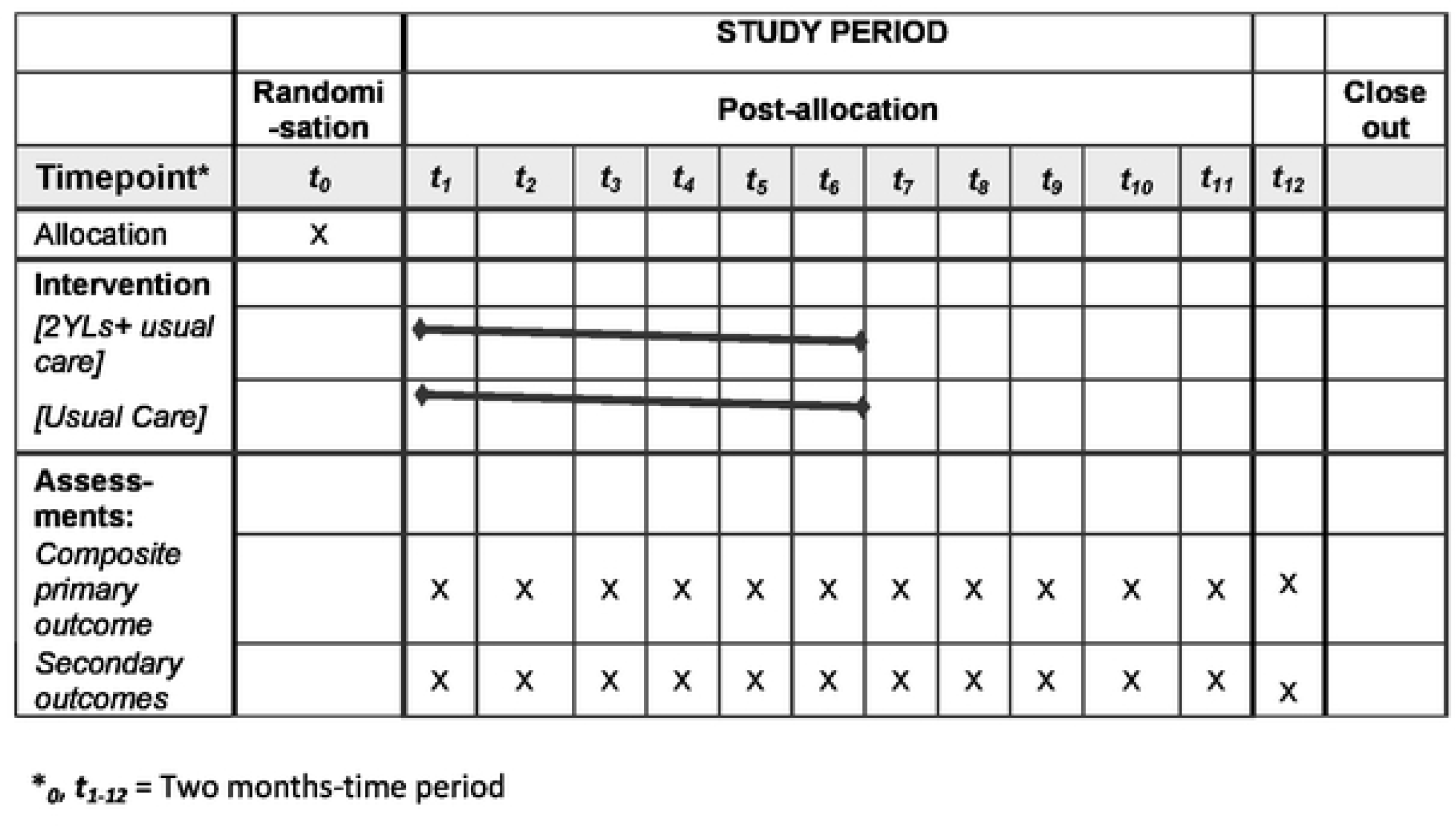

### Eligibility criteria

Eligibility criteria for the clusters is not having been previously exposed to the 2YL. All adolescent girls aged under 18 identified as pregnant in the community or presenting for maternity care at the selected clusters within the trial time frame will be included. There will be no exclusion criteria.

### Recruitment

Twelve clusters, (each with named communities routinely served by peripheral health units), will be identified by local partners and invited to participate in the study by the local research staff. Institutional-level consent and relevant local approvals from district health authorities and hospitals on behalf of the cluster will be obtained at the start of the trial (time point zero), and prior to intervention implementation. Participants living in the twelve clusters will be recruited by trained data collectors and can be identified through community members and different registers (i.e., antenatal, maternal and delivery register, referrals, outreach). In the event of changes to the PHUs serving a community during the study period (e.g. existing PHUs closing) we may invite a PHU to participate in the same randomised arm if it is serving the same community.

In intervention sites, pregnant adolescents can self-refer or be referred by a friend or family member who has heard about the mentoring scheme from community engagement activities. As per usual 2YL practice, the local team coordinator will meet with eligible pregnant girls and enrol them into the mentoring scheme as they come forward to a maximum of 36 (maximum number of girls that mentors can support per cluster due to limited resources). This helps us to test the feasibility of the intervention in a pragmatic way while avoiding selection bias. At their 6 weeks routine postnatal appointment, girls in both arms will be given the option to individually enrol and participate in two sub-studies (e.g. qualitative interviews, photovoice), and for those agreeing to participate in the sub-studies, explicit written informed consent (including from parents or guardians) will be gained and will be contacted around six-nine months after birth. Adolescents return to the PHU at one year for their routine infant immunisations appointment. A flow chart for 2YL is presented in Figure 2.

**Figure 2.**
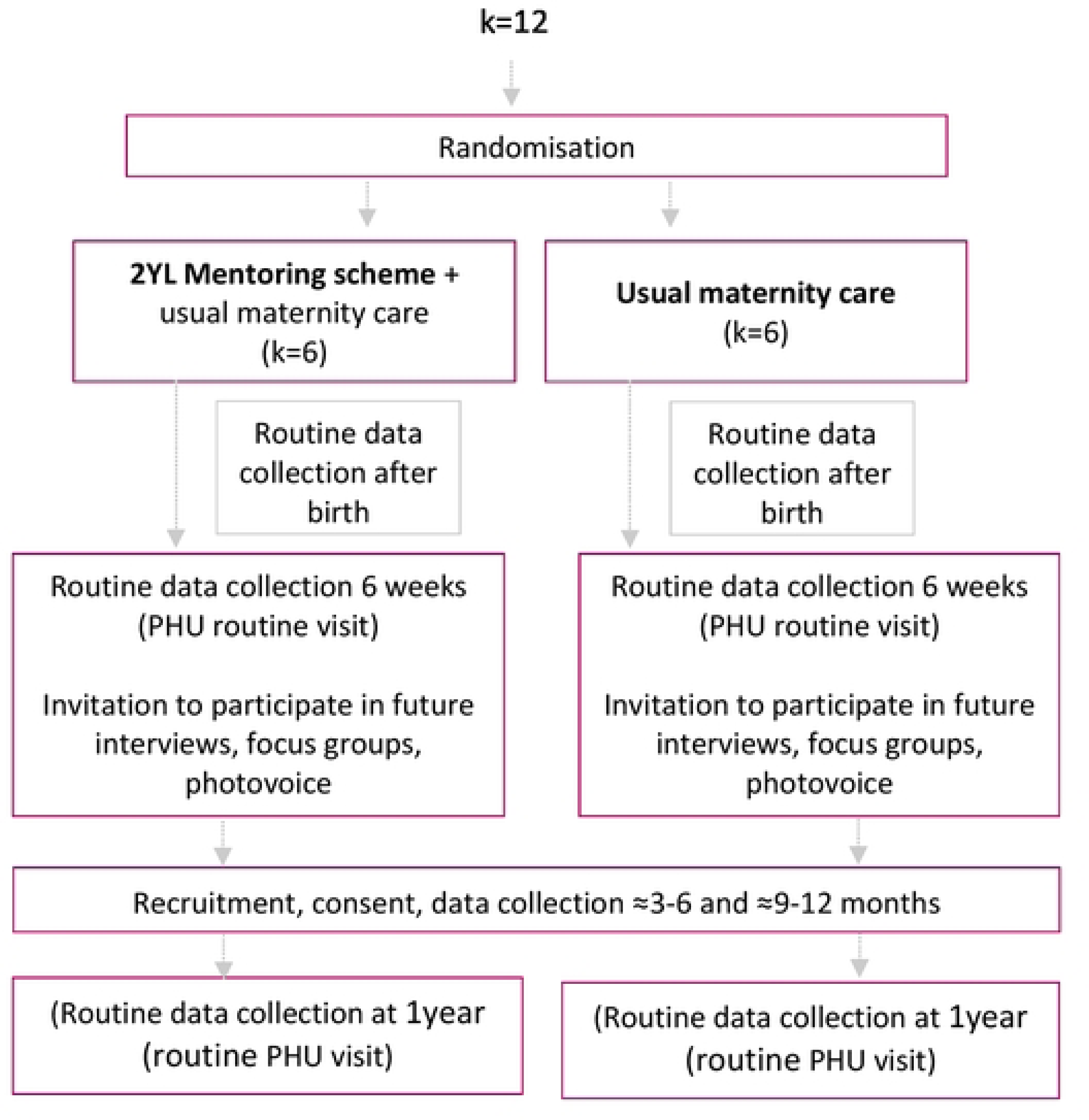
2YL pilot trial flowchart.

### Randomisation

The unit of randomisation is the trial area (or cluster), rather than the individual woman. The twelve clusters will be matched into pairs accounting for cluster size (births by adolescents) and distance from the referral hospitals. The clusters will be first allocated random cluster numbers and the sequence generation for receiving the 2YL intervention will then be determined by computer-generated random numbers by the trial statistician via a secure web-based randomisation and data management system hosted by MedSciNet with telephone back-up available at all times. The clusters will be masked until they are informed of their allocation two weeks prior to their implementation start date to give sufficient time for organizing community engagement activities, leading to recruitment and training of mentors and coordinators. A minimisation algorithm will also be used to ensure balance between groups with respect to PHU use and distance to referral hospital. The nature of the 2YL intervention is such that blinding of girls, mentors or healthcare providers cannot be achieved. However, outcome assessment will be masked to the 2YL statistician and the researchers who will analyse the data.

### Interventions

#### Intervention: 2YL + usual maternity care

Clusters and participants in the intervention group will receive the different components of the 2YL mentoring intervention (provided as an adjunct to usual maternity care). The structure of the 2YL intervention is focused saving adolescents’ lives and promoting their health and development.

Key roles: The intervention is run on the ground by a team from Lifeline Nehemiah Projects; with oversight from LNP’s Executive Director, a CEI specialist who leads on the initial community engagement activities. The project manager, a gender specialist, leads on recruitment, training and supervision of mentor teams, with support from a lead coordinator, and administrative and finance assistance from the LNP team.

Community engagement and involvement (CEI): this is a core first component of the 2YL intervention. LNP has devised a CEI strategy with three trips to each site to pave the way for the acceptance and tailoring of the intervention, considering local contextual factors, understanding how communities operate, and engaging those who would bring out various perspectives. Listening, discussing, and connecting with communities is imperative to build trusting relationships, and more CEI activities are undertaken in a bespoke fashion, depending on the specific requirements of each community.

Recruitment and training of mentors: women with a passion to support vulnerable girls are identified during CEI activities and recruited as mentors in collaboration with community stakeholders based on their experience, availability, knowledge of the community, commitment to improve girls’ lives and for having a reputation for kindness and being trustworthy. Each team also has a local coordinator who has the additional skill of literacy and record keeping allowing her to record details of activities and feed back to the central LNP team. Each site is scheduled to have a team of four mentors (and 1 local coordinator). Training and supervision: mentors and coordinators receive a 4-day manualised training programme using discussion, role play and bespoke pictorial maternal and newborn health knowledge and resources to learn and share important health messages and give practical support. The training emphasises the importance of commitment and confidentiality and is provided by an experienced educationalist with experience of training community members.

Ongoing support and supervision are provided to mentors by both their local coordinators and the LNP’s 2YL central management team. Mentors and coordinators are all volunteers who receive a monthly stipend to cover out of pocket expenses.

Matching of mentor-mentees: Adolescent girls are recruited to 2YL at any stage of pregnancy, and receive the mentoring for one year after birth (regardless of pregnancy outcome). They are identified by mentors in their localities, often by the girl herself hearing about the mentoring scheme from a peer, and approaching the mentor, or by the mentor hearing of a teenage pregnancy and asking the girl if she would like to join the scheme. Geographical proximity and shared tribal language are taken in consideration for matching girls with mentors. A total of 3-4 girls are matched to one mentor who will work with the adolescent for the entire one-year period unless covering or changing a mentor if needed for any arising issues.

2YoungLives activities include: at least weekly face to face meetings between mentor and mentee where the mentee feels comfortable and confidential conversation is possible; promotion of health services uptake; reminding to attend or attending with mentee for usual antenatal care; accompanying mentee to health facility when in labour or needing emergency care (or ensuring other birth partner is available according to mentee’s wishes); visits from mentor to mentee’s family to advocate for family support, if safe and appropriate; flexible and practical support for pregnancy and parenthood dependent on mentee’s support network; discussion with mentee to consider small business options and accompanying mentee to purchase first supply of goods; encouragement to return to school or start vocational training; a practical session making healthy baby food; encouragement to take up post-partum contraception; the importance of first line home treatment and early health-seeking behaviour for their baby; and an informal ‘graduation’ celebration at the end of the mentoring scheme. They also run monthly site meetings in the community with all mentors and mentees for peer support, cooking, eating and group discussion. At each of these sessions, visitors such as healthcare providers from local health facilities, teachers, other community members are invited for discussions about health topics or educational or training opportunities.

There are some aspects of the intervention that can be tailored depending on individual needs and circumstances; for example, if mentees are identified who have physical or intellectual disability, the team can enhance support with more visits and provide communication cards (for example for a deaf girl to use in labour) or by monitoring uptake of routine pregnancy medication (for girl with a learning disability). The whole team will always follow LNP’s safeguarding policies.

Usual or routine maternity care is described below and will be the same in both intervention and control groups.

#### Control: usual maternity care

The comprehensive care package will follow local and national guidelines for maternity care in Sierra Leone [20, 21]. Intervention packages prioritised under antenatal care are aligned to the 2016 WHO Antenatal model that recommends a minimum of eight antenatal contacts, with an overarching aim of providing pregnant adolescents or women with quality, respectful, individualised, person-centred care [22]. The latest Sierra Leone National Reproductive, Maternal, Newborn, Child and Adolescent Health Strategy outlines 1) interventions to be delivered under the antenatal care for positive pregnancy across the different service delivery levels, and 2) the priority for skilled birth attendance and essential newborn care all packaged under emergency obstetric and newborn care (EmONC) [20].

### Outcomes

#### Primary feasibility outcomes

Eligibility, recruitment, retention and attrition rates

Data collection and data completeness

Selection of most appropriate primary outcome measure and sample size calculation

#### Primary clinical outcome

The primary clinical outcome is a composite of maternal death (all-cause, occurring during pregnancy, labour, or within 42 days of birth), stillbirth (born with no signs of life at or after 28 weeks of pregnancy, but before or during birth) and neonatal death (deaths among live births during the first 28 days). We will report the effect of the 2YL intervention on both the composite and its components for all woman/baby deaths in the study; those who experience any one of the components will be considered to have experienced the composite outcome. Evaluating the data collection strategy and tool is an important element of testing the feasibility of a larger trial; for example, whether collecting outcome data up to six weeks postnatally is possible due to the mobility of this population.

#### Secondary clinical and process outcomes

Secondary maternal outcomes will include intermittent preventive therapy for malaria, anaemia, tetanus toxoid immunizations, caesarean sections, blood transfusions, birth at facility, sepsis, postpartum haemorrhage, place of birth and attendant, and uptake of contraception. Secondary perinatal outcomes will include; gestational age, birthweight, Apgar score, resuscitation, immediate breastfeeding, Kangaroo mother care, six weeks postnatal immunisation uptake, infant mortality.

Other secondary process and implementation outcomes will include service use, processes and implementation outcomes including; antenatal checks, antenatal checks with blood pressure measurement, referrals, postnatal check within a week of birth; measures of fidelity (e.g., mentors and team coordinators recruited, training packages and workshops, weekly one to one meetings, monthly social gatherings), acceptability (e.g. satisfaction with different components of the intervention among young girls, mentors, healthcare providers and local stakeholders), reach, adoption, women’s experiences of mentoring and care (e.g., respectful care access, trust, system responsiveness, quality and safety, attitude to and uptake of contraception in the postnatal year) *and* economic and social progress/wellbeing measures (e.g., return to education, experiences of running small businesses, perspectives of thriving), mechanisms of change (e.g., access to care, advocacy, support, respectful care, engagement, referral and escalation, health promotion, empowerment, relationships) and for social and economic wellbeing (e.g., social and economic empowerment, self-confidence, sense of agency, trustworthy adult and peers, respectful community engagement with families).

### Sample size

Sample size estimation has been provided by the 2YL trial statistician. Based on experience with some of the proposed trial areas and global maternal health literature relevant to these settings [4–7], we estimate the risk to adolescent pregnant women in Sierra Leone of experiencing a primary outcome (one of maternal death or stillbirth or neonatal death with no double counting) to be approximately 20% of all adolescent women. We also estimate a modest intra-cluster correlation (ICC) of 0.02 (sites similar but not identical, and a simple exchangeable autocorrelation structure based on previous experience in Sierra Leone) [23].

Prior to full proposal development, the 2YL team visited communities served by a typical PHU similar in size and distance to referral hospital to trial sites. At least 16% of all deliveries among teens had resulted in maternal death, stillbirth, or early neonatal death. It was not possible to include late neonatal deaths up to 28 days. In addition, girls who were referred to a referral hospital in an emergency were not included in these numbers. We believe that this data could match the indicative estimate of 20% of girls experiencing a primary outcome.

This study is planned as a pilot, with the main stated aim to demonstrate the feasibility of a fully powered study, and to help with its design. But for the purpose of the power calculation (intention to treat and not per protocol), an assumption of at least 42 deliveries per site has been made, with 20% having primary outcome composite events during the trial duration. These numbers will provide 84% power to detect a 52% relative risk reduction (RRR) of the primary outcome (from 20% to 9%) assuming a models ICC 0.02.

### Data collection

#### Primary and secondary pregnancy outcome

Primary and secondary pregnancy outcome data will be recorded over the study period and will be collected at an individual level from study-specific and routine data collection tools (e.g., antenatal attendance register, birth register, referral and outreach registers, 6-week postnatal check appointment book, hospital registers). At some PHUs, an additional ‘adolescent book’ is in use to record ANC registration of under-18-year-olds and associated birth outcomes. Data collection at both intervention and control clusters will be done by local data collectors who will enter the data anonymised onto MedSciNet, the study-specific management system.

#### Process Evaluation: experiences and implementation

In line with the current best practice in implementation research, the team will undertake a process evaluation to understand variations in the impact of the intervention on outcomes of interest and to contextualise findings. This will draw on the six core elements considered in the updated NIHR/MRC framework for evaluating complex interventions which include: interactions between intervention and context, theorising how the intervention may work, diversity of stakeholder’s perspectives in research, uncertainties, refinement of the intervention and resource and outcome consequences of the intervention [15]. A mixed methods approach will be used to assess implementation outcomes and identification of mechanisms and contextual factors that may influence variation in outcomes and experiences across multiple stakeholder groups, and at different stages of implementation. This offers a 360-degree evaluation, which considers needs and perspectives that typically differ between stakeholder groups and can vary over time.

Both quantitative and qualitative data on processes, experiences and implementation will be collected from monthly checklists, separate focus groups with a purposive sample of mentors, PHU healthcare providers and community stakeholders, and semi-structured interviews with some HCPs, community leaders, regional and national stakeholders, and adolescent girls in control and intervention sites. If appropriate, a family member or a friend will also be invited to participate in an interview. Girls will be offered the opportunity to be one-to-one (interviewer and participant) or one-to-two (interviewer and participant with chosen family members/friend). Thus, if appropriate, the family member/friend can be invited to the interviews, as social support is culturally patterned and could play an important part in the mechanisms of the intervention, particularly in women’s engagement with care. Wellbeing measures will include, for example, the adolescent flourishing scale [24] and a girl’s empowerment tool based on existing guides for impact evaluations [25]. A small sub-group of girls will also be invited to take part in the photovoice project, a participatory research methodology developed by Wang and Burrit [26] that partners with participants to use photography to help them to record, reflect upon, critically dialogue and share knowledge about their perspectives and priorities to reach policy makers, foster social change and strengthen partnerships [27].

### Statistical analysis

We will undertake two statistical analyses: the main pragmatic, intention to treat analysis (ITT) to compare intervention and control clusters that include all adolescent girls as originally allocated after randomisation; and a complementary per protocol (PP) analysis to compare intervention and control clusters that includes only those girls who received the intervention originally allocated. The CONSORT guidelines for reporting of parallel group RCTs recommend that both ITT and PP analyses should be reported for all planned outcomes to allow readers to interpret the effect of an intervention. It is not possible to blind clusters to interventions they receive, but outcome assessments will be blinded.

The main analysis will use a permutation test designed for stepped-wedge trials [28] to adjust the standard errors, confidence intervals and significance tests for clustering. This gives equal weight to each woman included in the study. We will adjust for important baseline differences that might be related to outcomes (e.g. parity, gestation at first antenatal visits). As individual-specific data will be used there is a potential for missing data; but the main impact of missing data would be to invalidate the randomised comparison if there was unequal dropout between the two arms, leading to potential bias. Multiple regression, as described, would correct for any such bias.

Qualitative data from interviews and focus group discussions will be mainly analyzed using thematic analysis [29]. In brief, this includes familiarisation with the data, generation of initial codes, the searching for and review of themes, naming and offering explanations for each theme, and lastly producing a report. This practical analytic approach involves inductive coding practices which are both consultative and initially open, and thus helpful to explore the perspectives of different research participants, highlighting similarities and differences, and generating unanticipated insights [29]. Implementation and contextual factors (barriers and facilitators) will be further assessed using the Consolidated Framework for Implementation Research [30] and Proctor’s framework [31,32]. A secondary grounded theory analysis will be undertaken to generate theory to explore and explain the phenomenon of adolescent pregnancy in Sierra Leone [33].

Implementation costs are important in process evaluations, and we will assess the feasibility of health economics for 2YL. It would not be appropriate to conduct a full economic evaluation for a pilot study designed to estimate the parameters needed to design a future definitive trial. Thus, the focus of health economics in this pilot trial will be limited to developing or refining service use schedules and other measures of outcomes. We will explore the resource use associated with the mentoring scheme and savings or other service use impacts and will also estimate costs associated with the delivery of the mentoring scheme. Cost consequences analyses will be used to present the results from this pilot study, as the costs and effects observed can also be used in value of information modelling to determine whether the cost of a definitive trial is worthwhile.

### Data management and monitoring

Anonymised data will be collected by the local data collectors, under the supervision of the trial manager. All participants will be given a unique identifier and no personal information will be entered onto the secure, online trial database (MedSciNet). Where possible, all anonymised data will be collected directly onto MedSciNet, but if exceptional circumstances make this not possible (e.g. no internet and very remote PHUs), a paper-based data collection tool will be used and stored anonymously in secure areas of each health facility and of the implementation partners. All data uploaded on the MedSciNet database will be automatically stored and backed up. Data collection and storage will be governed by the UK Data Protection Act 1998. All MedSciNet data is stored on high-capacity servers kept in highly secure monitoring and surveillance systems.

The MedSciNet database will be accessible for at least one year following the end of the trial, and a copy of this will then be kept on the KCL Server for 20 years, in accordance with the KCL Data retention schedule. Informed consent forms for qualitative components of the trial will be kept in files in secure areas of the management team (Lifeline Nehemiah Projects). Only the research assistant and the local and UK based managers will have access to these. All paper forms will be stored securely and kept in confidence in compliance with the UK Data Protection Act 1998.

The local research team will work in collaboration with the local community workforce and current local initiatives, to avoid missing primary outcome events. A flexible approach will be used to individualize our methodology according to local capacity and community healthcare workforce infrastructure. Consistency and quality of source data will be monitored by a research assistant and the Chief Investigators. MedSciNet allows for extensive monitoring and query processing features, as well as a comprehensive alerting system to identify missing data. Fields including ‘limits’ will be used to avoid entry of erroneous data. The 2YL management team will regularly conduct data monitoring visits to perform validation checks to verify validity and completeness of at least 10% of the source data. The team will also review the audit trail at individual levels if necessary. Training in the trial protocol will be delivered centrally (before trial recruitment) and on site in the field, to ensure local research staff are confident and competent to collect outcome data and ensure its quality, accuracy, and completeness. The final datasets used and/or analysed during the study will be made available from the corresponding author upon reasonable request.

### Trial oversight

This study will be supported by a trial management group (TMG) that includes research fellows, research assistants, data collectors, a 2YL programme manager and trial coordinator who will run the implementation and provide organisational support, particularly to the data collectors and teams in the field. The TMG will meet every week, with all co-investigators meeting every two-three months to monitor and review progress, troubleshoot and for strategic planning. This TMG reports to the steering committee, which is part of a wider international advisory group that oversees all activities of the different workstreams of the global health research group. The group will include dependent and independent members who will be invited to attend, at least, annual oversight meetings.

### Dissemination

Findings, positive or negative, will be disseminated in international peer-reviewed journals, as well as local, national and international events, workshops and conferences. Whenever possible, girls and their mentors, healthcare providers and community members and stakeholders will be provided with a layperson’s summary of the results, and government and policy makers will also be informed.

## Discussion

Sierra Leone has one of the highest maternal mortality rates in the world (717 deaths per 100 000 livebirths in 2019), and this burden falls primarily on adolescent girls. They are 40-60% more likely to die during childbirth and their infants are also at increased risk of sickness and death [1–2]. More than two thirds of all maternal deaths are caused by haemorrhage, hypertension and sepsis, and about a third are due to unsafe abortions among adolescents [2]. Many of these deaths could be prevented with simple, cost-effective and available interventions, but unfortunately there are often inequities in availability, access and quality of care, and delays in delivery, escalation and referral in Sierra Leone [14]. Adolescent girls are a particularly vulnerable group, often from disadvantaged communities and usually driven by poverty, lack of education and employment opportunities [1,2]. Stigma and abandonment, lack of family-based support and delayed care-seeking have been found to be important contributors to maternal adolescent mortality [4]. Social complexities and high rates of child marriage, adolescent pregnancy and gender-based violence (39%, 28%, and ∼50% respectively) also prevent girls from realizing their full potential in all aspects of their development [34,35].

The country has a very young population (nearly 65% are under 25 years old) [36], and improving health and wellbeing of these girls (including sexual and reproductive health) remains a top priority, indicated by a number of government policies targeted to this group: the Free Health Care Initiative for pregnant and breastfeeding women (2010), the National Standards for Adolescent and Young People Friendly Health Services (2011), the Reproductive, Maternal, Newborn, Child and Adolescent Health Policy (2017-2021); the National strategy for the reduction of adolescent pregnancy and child marriage 2018-2022 (2018), and the National Policy for Radical Inclusion in Schools (2021). The development of the first multi-agency and cross-ministry National Strategy for the Reduction of Teenage Pregnancy (2013-2015) was impeded by the scarcity of resources and diversion of efforts to the Ebola epidemic which also increased rates of adolescent pregnancy. However, in the aftermath of the epidemic, a revision and update of the national strategy was relaunched in 2018 which included child marriage [37].

Much of the attention has been given to primary prevention of adolescent pregnancy and child marriage, but adolescent girls are still getting pregnant and dying during childbirth. It is imperative to also identify programmes or interventions that support them during pregnancy and the parenting period so they can survive and improve their health and development and that of their child. To support and facilitate the integration of pregnant and parenting girls it is crucial to sensitise communities, strengthen existing youth friendly services and work closely with stakeholders (government, gatekeepers, community and family actors). It is now widely recognized that the need to support a holistic agenda where integration and implementation of evidence-based interventions across health, education, and social systems must improve in order to protect, nurture, and support the health and development potential for every child and adolescent [38].

These are the philosophical principles by which the 2YL community-based mentoring intervention was initially developed in 2017. Mentored adolescent girls are supported to thrive, not just survive. Youth mentoring programmes have been shown to improve outcomes across academic, behavioral, emotional and social areas of young people’s lives. At present there is no clear evidence that they can improve physical health, although studies are limited (including for pregnant and parenting adolescents) [39,40]. The evidence suggests that longer mentoring relationships are linked to better outcomes, and the role of the matching process, the training and motivation, and the need for goal-orientated programmes are keys for success [39]. Goldner and Ben-Eliyahu [41] tried to unpack the relational processes in community-based youth mentoring which promote high relationship quality and generate the most significant benefits. They found that long enough, supportive, reliable, trustworthy, and balanced mentoring relationships (clearly articulated goals, structure, and behaviours) serve as building blocks in promoting mentees’ development and minimising adversity. In addition, consideration needs to be given to moderating factors (i.e. mentors’, mentees’ and matching characteristics), highlighting the need for more research in this area [41]. Although the 2YL mentoring scheme has been piloted in five sites over four years with promising results, a more robust and formal evaluation is needed to understand the feasibility of 2YL in other communities and how it can address determinants of adolescent maternal mortality and general health and wellbeing, and this is what the 2YL pilot trial is aiming to achieve.

Since 2021, 2YL has been part of a Global Health Research Group (CRIBS) funded by the UK’s National Institute for Health and Care Research and led by the University of Sierra Leone and King’s College London in collaboration with multiple partners, including the Sierra Leone Ministry of Health and Sanitation, Lifeline Nehemiah Projects (LNP), NGO Welbodi Partnership, the National Emergency Medical Services (NEMS), and the national Midwifery Schools. The group, which will run for 3 years, aims to develop and implement simple, scalable innovations to reduce maternal (including adolescent) and perinatal mortality and build research capacity and expertise in Sierra Leone [42]. The group is uniquely multidisciplinary with expertise in reproductive health, obstetrics and gynecology, nursing, midwifery, public health, gender studies, sociology, implementation science, medical statistics, health policy and economics.

We believe findings from the 2YL pilot trial will provide more holistic information for communities and local and national decision-makers and refine procedures to inform future scale-up work aiming to reduce mortality among adolescent girls and their babies, and improving their health, educational and socio-economic welfare. The various programmes implemented by the line ministries in Sierra Leone are more institutionally based with little active involvement with communities that would serve to support prevention and sustainability; one of the unique features of the 2YL model is the meaningful community engagement and involvement. There has been national and international interest in 2YL which has been featured in the Lancet [10], the 2022 UNFPA State of the World Population Report [43], a programme for the BBC World Service ‘People fixing the World’ [44] and the Evening Standard [45]. We believe 2YL has the potential to save lives and promote health and wellbeing of adolescent girls and babies, and thus become a model of good practice for adolescent pregnancy, to be adopted more widely in Sierra Leone and elsewhere.

## Supplementary information

No data are available.

## Trial status

The current 2YL protocol is version 2.1, 21 July 2023. Recruitment start time was 4 July 2022; completion is 30 November 2023.

## Abbreviations

2YL: 2YoungLives
cRCT: cluster randomised controlled trial
CONSORT: Consolidated Standards of Reporting Trials
EmONC: Emergency Obstetric and Newborn care
ICC: Intra-Cluster Correlation
ISRCTN: International Standard Randomised Controlled Trial Number
KCL: King’s College London
PHU: Peripheral Health Unit
SL: Sierra Leone
SPIRIT: Standard Protocol Items: Recommendations for Interventional Trials
TMG: Trial Management Group
TIDieR: Template for Intervention Description and Replication
WHO: World Health Organization.

## Declarations

### Availability of data and Material

No data are available yet.

### Authors’ contributions

LN, MK, PTW are the co-founders of 2YL. CFT, LN, MK drafted the initial trial protocol, and PK, AMK, AR, ST, PTS, AHS, JS and PTW reviewed and commented on the initial draft and/or on subsequent revisions. All authors have seen and approved the final version of the manuscript.

### Ethical approval and consent to participate

This study has been approved by ethics committee at King’s College London UK (HR/DP-21/22-26320) and the Office of the Sierra Leone Ethics and Scientific Review Committee. Data security will be maintained in accordance with General Data Protection Regulations. Written, informed consent to participate will be obtained from all participants for qualitative interviews, focus group discussions, and photovoice.

### Consent to publication

This manuscript does not contain individual personal data from participants.

## Acknowledgements

We thank all members of CRIBS Group & collaborators at the University of Sierra Leone, King’s College London; Ministry of Health and Sanitation; Schools of Midwifery; Welbodi Partnership; NEMS; PCMH; UNICEF; WHO SL. Special thank you to CRIBS research assistants and data collectors working for 2YL, as well as all the girls, mentors, relatives, healthcare providers and community stakeholders.

## Name and contact information for sponsor

King’s College London (Professor Reza Razavi)

## References

1. WHO, UNICEF, UN Population Fund, World Bank Group and the United Nations Population Division. Maternal mortality: levels and trends 2000 to 2017. Geneva: World Health Organization, 2019.

2. Maternal death surveillance and response Annual Report. A call for action, time to respond. Ministry of Health & Sanitation, Directorate of Reproductive and Child Health, Sierra Leone. 2019.

3. UNFPA, National strategy for the reduction of adolescent pregnancy and child marriage, 2018-2022.https://reliefweb.int/sites/reliefweb.int/files/resources/SLE_country%20profile.pdf (accessed Feb 22, 2022).

4. November L, Sandall J. ‘Just because she’s young, it doesn’t mean she has to die’: exploring the contributing factors to high maternal mortality in adolescents in Eastern Freetown; a qualitative study. Reprod Health 2018; 15: 31.

5. Vogel JP, Pileggi-Castro C, Chandra-Mouli V, et al. Millennium development goal 5 and adolescents: looking back, moving forward. Arch Dis Child 2015;100 Suppl 1:S43–7

6. Nove A, Matthews Z, Neal S, et al. Maternal mortality in adolescents compared with women of other ages: evidence from 144 countries. Lancet Glob Health 2014;2:e155–64.

7. Li, Zhihui, et al. “Maternal healthcare coverage for first pregnancies in adolescent girls: a systematic comparison with adult mothers in household surveys across 105 countries, 2000– 2019.” BMJ Global Health 5.10 (2020): e002373.

8. https://2younglives.org/

9. Kamara M, November L. 2 Young Lives: mentoring teenagers for safer pregnancy and birth; Project report 2020. 2 Young Lives website. June 2020. Accessed October 05, 2021. https://2younglives.org/wp-content/uploads/2020/07/2YL-2020-report-final.pdf

10. Fernandez Turienzo C, November L, Kamara M, Kamara P, Goodhart V, Ridout A. et al. Innovations to reduce maternal mortality and improve health and wellbeing of adolescent girls and their babies in Sierra Leone. The Lancet Child & Adolescent Health. 2023 Mar 1;7(3):151–3.

11. ICF. SSL (Stats S and. Sierra Leone. Sierra Leone Demogr Heal Surv 2019. 2019;9:187–93.

12. BBC World. Sierra Leone declares emergency over rape and sexual assault. Available from: https://www.bbc.co.uk/news/world-africa-47169729 (Accessed 19 September 2023)

13. The National health Workforce Accounts database, World Health Organization, Geneva 2022.Available from: https://apps.who.int/nhwaportal, https://www.who.int/activities/improving-health-workforce-data-and-evidence

14. UNICEF. Maternal and newborn health disparities, Sierra Leone. https://reliefweb.int/sites/reliefweb.int/files/resources/SLE_country%20profile.pdf (accessed June 22, 2023).

15. Skivington K, Matthews L, Simpson SA, Craig P, Baird J, Blazeby JM, et al. A new framework for developing and evaluating complex interventions: update of Medical Research Council guidance. bmj. 2021 Sep 30;374.

16. Curran GM, Bauer M, Mittman B, Pyne JM, Stetler C. Effectiveness-implementation hybrid designs: combining elements of clinical effectiveness and implementation research to enhance public health impact. Medical care. 2012 Mar;50(3):217.

17. Eldridge SM, Chan CL, Campbell MJ, Bond CM, Hopewell S, Thabane L, Lancaster GA. CONSORT 2010 statement: extension to randomised pilot and feasibility trials. bmj. 2016 Oct 24;355.

18. Chan AW, Tetzlaff JM, Gøtzsche PC, Altman DG, Mann H, Berlin JA, et al.. SPIRIT 2013 explanation and elaboration: guidance for protocols of clinical trials. Bmj. 2013 Jan 9;346.

19. Hoffmann T, Glasziou P, Boutron I, Milne R, Perera R, Moher D, Altman D, Barbour V, Macdonald H, Johnston M, Lamb S, Dixon-Woods M, McCulloch P, Wyatt J, Chan A, Michie S. Better reporting of interventions: template for intervention description and replication (TIDieR) checklist and guide. BMJ. 2014;348:g1687.

20. Ministry of Health and Sanitation. Sierra Leone National Reproductive, Maternal, Newborn, Child and Adolescent Health Strategy 2017–2021. Sierra Leone: Ministry of Health and Sanitation. 2017.

21. Ministry of Health and Sanitation. Sierra Leone basic package of essential health services 2015-2020.

22. . World Health Organisation. WHO recommendations on antenatal care for a positive pregnancy experience. Geneva: WHO, 2016

23. Vousden N, Lawley E, Nathan HL, Seed PT, Gidiri MF, Goudar S, Sandall J, Chappell LC, Shennan AH, Kachinjika M, Bukani D. Effect of a novel vital sign device on maternal mortality and morbidity in low-resource settings: a pragmatic, stepped-wedge, cluster-randomised controlled trial. The Lancet Global Health. 2019 Mar 1;7(3):e347–56.

24. Waigel NC, Lemos VN. A systematic review of adolescent flourishing. Europe’s Journal of Psychology. 2023 Feb;19(1):79.

25. Glennerster R, Walsh C, Diaz-Martin L. A practical guide to measuring women’s and girls’ empowerment in impact evaluations. Gender Sector, Abdul Latif Jameel Poverty Action Lab. 2018 Nov 28.

26. Wang C, Burris MA. Photovoice: concept, methodology, and use for participatory needs assessment. Heal Educ Behav. 1997;24:369–87.

27. Budig K, Diez J, Conde P, Sastre M, Hernán M, Franco M. Photovoice and empowerment: evaluating the transformative potential of a participatory action research project. BMC public health. 2018 Dec;18(1):1–9.

28. Thompson J, Davey C, Hayes R, Hargreaves J, Fielding K. Permutation tests for stepped-wedge cluster-randomized trials. The Stata Journal. 2019;19(4):803–19.

29. Braun V, Clarke V. Thematic analysis. American Psychological Association; 2012.

30. Damschroder LJ, Reardon CM, Widerquist MA, Lowery J. The updated Consolidated Framework for Implementation Research based on user feedback. Implementation science. 2022 Dec;17(1):1–6.

31. Proctor E, Silmere H, Raghavan R, Hovmand P, Aarons G, Bunger A, Griffey R, Hensley M. Outcomes for implementation research: conceptual distinctions, measurement challenges, and research agenda. Administration and policy in mental health and mental health services research. 2011 Mar;38:65–76.

32. Proctor EK, Bunger AC, Lengnick-Hall R, Gerke DR, Martin JK, Phillips RJ, Swanson JC. Ten years of implementation outcomes research: a scoping review. Implementation Science. 2023 Dec;18(1):1–9.

33. Strauss A and Corbin. Basics of qualitative research: Grounded theory procedures and technique. 2nd ed. Newbury Park: Sage, 1998

34. UNICEF. Ending child marriage and teenage pregnancy in Sierra Leone. https://reliefweb.int/sites/reliefweb.int/files/resources/SLE_country%20profile.pdf (accessed June 22, 2022).

35. UN Population Fund. Country profile Sierra Leone. https://reliefweb.int/sites/reliefweb.int/files/resources/SLE_country%20profile.pdf (accessed September 12, 2023).

36. Sierra Leone 2015 statistics child, adolescent and youth report https://www.statistics.sl/images/StatisticsSL/Documents/Census/2015/sl_2015_phc_thematic_report_on_children_adolescents_and_youth.pdf (accessed June 22, 2022).

37. Ministry of Health and Sanitation, Government of Sierra Leone. National Strategy for the reduction of adolescent pregnancy and child marriage 2018-2022. Government of Sierra Leone, 2018.

38. Bhutta ZA, Boerma T, Black MM, Victora CG, Kruk ME, Black RE. Optimising child and adolescent health and de

39. Raposa EB, Rhodes J, Stams GJ, Card N, Burton S, Schwartz S, Sykes LA, Kanchewa S, Kupersmidt J, Hussain S. The effects of youth mentoring programs: A meta-analysis of outcome studies. Journal of youth and adolescence. 2019 Mar 15;48:423–43.

40. Armitage H, Heyes K, O’Leary C, Tarrega M, Taylor-Collins E. (2020) What Makes for Effective Youth Mentoring Programmes: A rapid evidence summary. Nesta: Manchester Metropolitan University.

41. Goldner L, Ben-Eliyahu A. Unpacking community-based youth mentoring relationships: An integrative review. International journal of environmental research and public health. 2021 May 25;18(11):5666.

42. https://cribs-i.org (Accessed 14 September 2023)

43. UNFPA. State of World Population 2022: Seeing the Unseen: The Case for Action in the Neglected Crisis of Unintended Pregnancy (Page 49) Available from: https://www.unfpa.org/sites/default/files/pub-pdf/EN_SWP22%20report_0.pdf (Accessed 14 September 2023)

44. BBC World Service ‘People fixing the World’’ Helping teenagers become good mums. Available from: https://www.bbc.co.uk/programmes/p0c8jwj8 (Accessed 14 September 2023)

45. Evening Standard: Sierra Leone’s pregnant teens matched with mentors to save their lives. Available from https://www.standard.co.uk/optimist/let-girls-learn/sierra-leone-pregnant-teenagers-sex-mentors-childbirth-education-b1058657.html (Accessed 14 September 2023)

